# Optimizing Trastuzumab Duration: Cost-Effectiveness Analysis of Five Regimens for HER2-Positive Breast Cancer in Kenya

**DOI:** 10.64898/2026.05.12.26353063

**Authors:** Prashant Mandaliya, Edwine Barasa, Dorothy Aywak, Faith Okalebo

## Abstract

Breast cancer was the leading cause of cancer-related mortality among women worldwide in 2022. In Kenya, more than a quarter of breast cancer patients have the aggressive Human Epidermal Growth Factor Receptor 2 positive subtype. Trastuzumab is recommended for its treatment, but high costs have limited access. This study evaluated the cost-effectiveness and affordability of trastuzumab-based regimens to inform their adoption and use in Kenya.

A cost-utility analysis was conducted from the healthcare payer perspective over a lifetime horizon. Five trastuzumab-based regimens of varying durations (9-week, 6-month, 9-month, 12-month, and 24-month) were compared with chemotherapy alone. Direct medical costs were estimated using a bottom-up micro-ingredient approach. All costs were reported in 2022 USD. A cohort Markov state-transition model with a monthly cycle length was used to estimate the costs and outcomes for an open hypothetical cohort. Scenario, deterministic sensitivity and probabilistic sensitivity analyses were conducted. A budget impact analysis estimated the financial implications of each regimen.

The 9-week regimen had the lowest incremental cost-effectiveness ratio (ICER) of USD 3,230 per QALY, while the remaining regimens had ICERs ranging from USD 4,046 to 9,846 per QALY. The findings were most sensitive to the price and quantity utilized per cycle of trastuzumab. A reimbursement cap of KES 40,000 per cycle reduced ICERs by up to 61%. Over five years, the 9-week regimen would account for 1.2% of the projected insurer’s budget, whereas the current recommended 12-month regimen would consume 2.82%.

Although none of the regimens were cost-effective at Kenya’s WTP threshold (USD 1054.80), the 9-week regimen may still be considered by policymakers given its greater affordability. Further cost reductions can be achieved through negotiating lower drug prices, improving access to biosimilars, and implementing vial sharing.

## Introduction

In 2022, breast cancer was the leading cause of cancer related mortality among women worldwide, and the second most prevalent malignancy [1]. Treatment outcomes for breast cancer are poorer in African countries, with a case-fatality rate of nearly 50% [2]. In Kenya, cancer incidence continues to rise, with 44,726 new cases and more than 100,000 prevalent cases recorded in 2022 [3]. Human epidermal growth factor receptor 2-positive (HER2+) breast cancer is a more aggressive form of the disease with over a quarter of breast cancer cases in Kenya testing HER2 positive [4].

Trastuzumab is a humanized monoclonal antibody used in the treatment of the disease as it targets the HER2 receptor. Large clinical trials have demonstrated the drug’s effectiveness, showing reductions in recurrence risk of up to 52% and mortality by 33%, when compared to trastuzumab-naïve regimens [5]. Evidence shows that initiating treatment with trastuzumab at an earlier stage of the disease is linked to better survival and treatment outcomes [6]. The American Society of Clinical Oncology (ASCO) and the US National Comprehensive Cancer Network (NCCN) recommend trastuzumab for the treatment of HER2+ breast cancer [7,8]. In Kenya, the National Cancer Treatment Protocol also recommends treatment with the drug, using the 12-month regimen [9]. Although shorter regimens have been shown to be effective in large clinical trials [10–12], research regarding their cost-effectiveness remains limited within the African context.

A cost-utility analysis (CUA) would provide policymakers with evidence on the cost-effectiveness of trastuzumab in the Kenyan context and help identify which regimen provides the best value for money. When integrated with a budget impact analysis (BIA), the analysis provides a more comprehensive understanding of the affordability and financial sustainability of the interventions. This evidence is particularly important in low and middle income countries (LMICs), where resources are scarce and access to high-cost treatments remains limited [13]. Out-of-pocket (OOP) payments for treatment with trastuzumab remains a barrier to treatment, with less than 5% of patients in Africa being able to afford treatment with the drug [14]. This has resulted in increased non-adherence and treatment delays, leading to worse outcomes. A recent study highlighted that survival rates of breast cancer patients in LMICs were nearly 25 times lower than those of patients from high-income countries, with access to effective treatment being one of the main contributors to this difference [15].

Historically the National Health Insurance Fund (NHIF), only reimbursed four trastuzumab cycles per year in Kenya, with a Kenya Shillings (KES) 150,000 cap per cycle, leaving patients to cover the remaining costs [16]. As of October 2024, the Social Health Authority (SHA) replaced NHIF as the national insurer with the aim to advancing universal health coverage [17]. While oncology related benefits were initially capped at KES 400,000 per year, they have now doubled to KES 800,000 under the expanded oncology care package [18,19]. This expansion was complemented with a landmark Memorandum of Understanding (MOU) signed in 2025 between the Ministry of Health and Roche Pharmaceuticals, which capped the cost of trastuzumab to KES 40,000 per cycle and eliminated the need for OOP co-payments for patients covered under the SHA framework [20].

Prior economic analyses from high-income countries have found trastuzumab to be cost-effective, with results heavily dependent on drug price, time horizon, and willingness-to-pay threshold [21]. To the best of our knowledge, there is only one multi-country study that has examined the cost-effectiveness of the 12-month trastuzumab regimen in Kenya [22]. However, the study used a single generalized estimate for the cost of treatment with trastuzumab across all included countries [22]. Furthermore the study only assessed the 12-month trastuzumab regimen, and did not conduct a budget impact analysis [22]. Given that costs are highly context-specific, these estimates may not be ideal for localized decision-making.

In cognizant of these gaps, this study aimed to provide contextual evidence on the cost-effectiveness and affordability of five different trastuzumab regimens in Kenya. Local cost data were utilized, and the analysis was conducted from the healthcare payer perspective. The outputs from the study can be used by members of the Benefits Package and Tariffs Advisory Panel (BPTAP), to advise SHA on the inclusion of trastuzumab in the national health benefits package for Kenya.

## Methodology

### Study design

The study was conducted in three distinct phases. The initial phase involved a bottom-up micro-costing analysis, which aimed to identify the direct medical costs associated with treatment of HER2+ breast cancer in Kenya. Next, a cost-utility analysis was conducted using a cohort Markov state-transition model to determine the incremental cost-effectiveness ratio (ICER) of each regimen when compared with the standard of care. Finally, a budget impact analysis was conducted to assess affordability and financial impact for the national insurer.

### Trastuzumab-based treatment regimens

The regime used as the reference group included four cycles of an anthracycline-containing agent and cyclophosphamide, followed by four cycles of paclitaxel. This regimen represents the most affordable chemotherapy option currently available for HER2+ breast cancer patients in Kenya.

Treatment regimens were selected based on a literature review of clinical evidence, incorporating all protocols with established effectiveness results. Four of the five selected trastuzumab-based interventions had the same non-biologic chemotherapy regimen as the reference group, which included four cycles of trastuzumab that were initiated concurrently with paclitaxel, followed by monotherapy with trastuzumab for a specified number of cycles, based on the total duration of the regimen.

The 9-week regimen followed a different approach, with paclitaxel first administered on its own for three cycles, followed by a combination of fluorouracil, epirubicin, and cyclophosphamide (FEC) for three cycles [10]. Finally, trastuzumab is administered as monotherapy for three cycles. The details for each regimen are described in Table 1.

**Table 1:** Trastuzumab regimens used for management of HER2 positive breast cancer.

| Name | Regimen | Reference |
| --- | --- | --- |
| Chemotherapy only (Reference group) | AC (4) + T (4) | Perez et al. [23] |
| 9-week regimen | T (3) + FEC (3) + H (3) | Joensuu et al. [10] |
| 6-month regimen | AC (4) + TH (4) + H (5) | Deng et al. [12] |
| 9-month regimen | AC (4) + TH (4) + H (9) | El-Enbaby et al. [11] |
| 12-month regimen | AC (4) + TH (4) + H (13) | Perez et al. [23] |
| 24-month regimen | AC (4) + TH (4) + H (30) | Cameron et al. [24] |

### Study perspective, time horizon and discount rate

The analysis was conducted from the healthcare payer perspective, incorporating all direct medical expenditures relevant to the Kenyan public health system. A lifetime horizon was adopted. All costs were reported in 2022 KES, and converted to 2022 USD using a conversion rate of USD 1 to KES 121 [25]. A discount rate of 6% was applied to costs and 4% for utilities, as suggested by Haacker et al. [26].

### Cost analysis

#### Study design

A cost analysis was conducted using a micro-costing and bottom-up approach. Data on costing items, which included the unit cost, probability of use, and quantity, were obtained from multiple sources, such as key informant interviews (KIIs), market surveys, treatment guidelines, and literature.

#### Study site

The study site for the KIIs was Kenyatta National Hospital (KNH). It is the largest public tertiary referral hospital in Kenya and is located in Nairobi. It also serves as the teaching hospital for the University of Nairobi [27]. Patients from across Kenya and parts of East/Central Africa are seen at KNH as they offer specialty diagnostic, preventive and curative health services. It is also one of the largest public providers of cancer related services in the country [27].

A market price survey was conducted in Nairobi County to obtain local prices for medicines and laboratory tests. Nairobi is the commercial capital of Kenya, and a majority of laboratories and pharmaceuticals suppliers have branches in the county.

#### Key informant interviews

Interviews were conducted with healthcare workers (n=18) of different cadres (oncologists, anesthesiologists, surgeons, radiotherapists, nurses and pharmacists) directly involved in the care of breast cancer patients at KNH from 1st August 2020 to 1st February 2021. Purposive sampling was used for the selection of the key informants. Participants were required to be healthcare workers directly involved in the care of breast cancer patients, with at least 2 years of employment at KNH, and provided verbally informed consent. The principle of saturation was used to determine the sample size [28]. Data were collected using a semi-structured, cadre-specific interview guide, containing questions on prices, quantities, and probabilities of resource use. Interviews were recorded and transcribed within 24 hours. All recordings were deleted once transcription was complete. The information obtained from each key informant was entered into a pre-designed MS Excel table to collate and organize the data.

#### Market survey

A market survey was conducted across established suppliers of pharmaceuticals and non-pharmaceutical products, as well as clinical laboratories. Universal sampling was utilized whereby all known suppliers for each cost item were approached, with a minimum of three price quotes obtained per item to ensure comparability. For pharmaceuticals that were only available as the originator brand, the price came from a single source due to limited suppliers. To account for price variability, prices for the lowest- and highest-priced generics and the originator brand were collected and used as inputs in the sensitivity analyses. Base prices were obtained from the finance department at KNH as it often serves as a benchmark for public sector tariffs.

The prices for laboratory tests and diagnostic imaging procedures were obtained through a survey of major laboratory service providers in Nairobi, which included KNH, two large private hospitals, and a specialist private laboratory. Additional hospital charges, such as those for consultation and bed fees, were obtained from KIIs and KNH.

#### Costs components excluded

Administrative expenses, capital and maintenance costs of equipment, and cleaning costs were excluded, as they were assumed to be similar across all treatment regimens. The cost of managing congestive heart failure, a potential side effect of trastuzumab, was excluded as KIIs indicated that the side effect was very rare in the Kenyan setting.

#### Computation of direct medical costs

Direct medical costs were estimated by accounting for all the resources utilized by a patient throughout their clinical journey. These costs were categorized as diagnostic, treatment, follow-up and palliative care-related resources. The subsequent sections describe the methodology used to estimate each cost component in detail.

#### Pharmaceuticals and non-pharmaceuticals

Quantities for pharmaceuticals were determined using data obtained from clinical protocols and KIIs. For regimens requiring body weight- or body surface area for dosing calculations, a mean body weight of 65.5 kg and height of 157.9 cm were utilized, as reported by a study on breast cancer patients in Kenya [29]. The quantities of any pharmaceutical present in a single dose formulation were rounded up to the next whole number. No sharing of pharmaceuticals between patients was assumed to ensure all wastages were captured as expenditures. The cost of non-pharmaceuticals utilized during administration of medication was captured through a standardized administration fee charged at KNH.

#### Diagnosis

The tunnel state representing “Diagnosis” included all the resources consumed in diagnosing a patient with breast cancer. This included consultation fees, laboratory tests and diagnostic procedures that are routinely requested for diagnosis. The total cost of each item was calculated by multiplying the probability of an item being used by the quantity consumed and its unit price.

#### Surgery

The cost of surgical procedures routinely performed in breast cancer patients (modified radical mastectomy and breast conservation surgery) were estimated. The cost of resources such as anesthesia, theater fees, nursing fees, inpatient charges and those for pharmaceuticals consumed both pre- and post-surgery were included. Based on insights from KIIs, a weighted average using the probabilities of undergoing each type of surgery was calculated to represent the total cost for surgery, while the lower and upper estimates were populated based on information provided in the KIIs. The baseline value for the total cost for surgery was USD 409.64 (Range: USD 165.29 - USD 2479.34)

#### Treatment, remission, and palliative states

The model included tunnel states for primary disease treatment, maintenance therapy during remission, and palliative care during metastasis. For each tunnel state, the total cost were estimated by summing all resources consumed which included medications, procedures, hospital services and adverse effect management. For items not consumed on a monthly basis, a probability of use was applied to ensure the valuation reflected the expected monthly expenditure rather than a fixed recurring fee. This approach was applied across treatment, remission and palliative care related tunnel states.

#### Five-year cost per treatment regimen

To examine how costs varied across years over a five-year costing time horizon, disease-free survival health states were developed for each treatment regimen arm using tunnel states to reflect their specific resource requirements. For each regimen arm, sixty monthly tunnel states were modelled within the disease-free survival health state, with costs assigned to each tunnel state reflecting the resource requirements at that point in the treatment pathway. Monthly costs were aggregated annually to illustrate how costs evolved over the five-year period, and total costs over the full five-year horizon were summed to enable comparison across regimens.

### Cost-utility analysis

#### Study design

The cost utility analysis was conducted using a Markov state-transition model to estimate incremental costs and health outcomes. The model utilized a monthly cycle length over a lifetime horizon with time dependent transition probabilities, reflecting changes in clinical risks over time.

#### Markov model

A discrete cohort state-transition Markov model with non-stationary (time-dependent) transition probabilities was utilized to capture the dynamic nature of breast cancer management. A one-month cycle length was used because treatment and disease progression can change rapidly in breast cancer, with monthly updates to costs and utility values, making the model non-homogeneous [30]. The model included the following five health states: disease-free survival, local or regional recurrence (LR), metastasis (Met), all-cause mortality (ACM), and death by breast cancer (DBC). All patients entered the model through the DFS state at the time of their first consultation with the oncologist during diagnosis. To capture the time-dependent changes, each state was implemented as a sequence of tunnel states, which were arranged chronologically from month 1 to month 60 for each health state in which patients were alive (disease free survival, local recurrence, and metastasis). These are detailed in S1_Appendix.

Patients that transitioned from local recurrence back to disease free survival joined the remission-related tunnel state to accurately reflect the clinical pathway of successfully treated recurrent cases. Those patients that reached the final tunnel state of a given health state remained in that tunnel state in subsequent cycles until they transition to a different health state. Transitions between the health states are illustrated in Fig1.

**Fig1:**
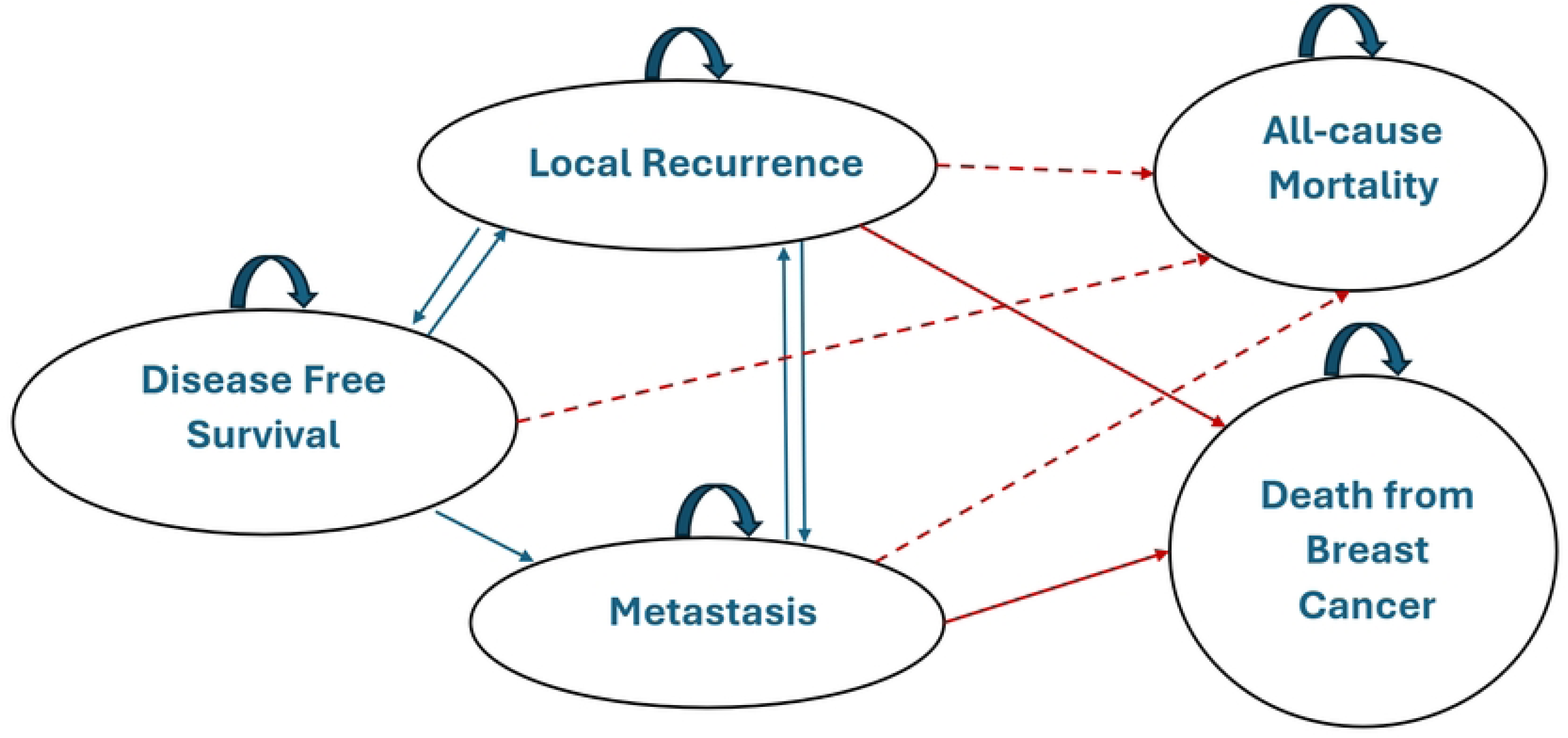
Markov transition states for breast cancer patients on chemotherapeutic regimens

An open cohort of hypothetical patients was used in this model, simulating an enrollment of 100 patients per month over 5 years, following an initial group of 100. As a result, 1,200 patients joined the cohort annually, resulting in a total of 6,100 patients initiating treatment over the five-year period, after which the cohort was closed. The decision to enroll 100 patients per month was informed by epidemiological data from Kenya, where approximately 6,000 new breast cancer cases were diagnosed in 2018 [31], with an estimated 25% classified as HER2+ [32]. The model was implemented in Base R (version 4.3.2) using the *dampack* package [33].

#### Model inputs

The movement of patients across the different health states was determined using transition probabilities. Data for calculating transition probabilities for the 12-month group were obtained from literature with a focus on LMICs, as Kenyan specific survival data was scarce. The only available Kenyan studies yielded implausibly low transition probabilities, largely due to significant loss to follow-up, small sample sizes, and short follow-up periods, which limited their reliability for modeling [34,35]. To account for background mortality, age-specific life expectancy was derived from WHO life tables for Kenya [36], allowing the probability of all-cause mortality to vary dynamically as the cohort aged within the model. Table 2 summarizes the monthly transition probabilities for the 12-month regimen.

**Table 2:** Monthly transition probabilities for the 12-month regimen.

| Parameter | Base value | 95% CI | Source |
| --- | --- | --- | --- |
| **Year 1** |  |  |  |
| DFS > LR | 0.00175 | 0.0015 - 0.0019 | Gupta et al. [37] |
| DFS > Met | 0.003 | 0.0026 - 0.0033 | Gupta et al. [37] |
| LR > DFS* | 0.0088 | 0.0069 – 0.0106 | Estimated based on 10% salvage rate. |
| LR > Met** | 0.00937 | 0.00746–0.01148 | Elsisi et al. [38] |
| LR > DBC | 0.0079 | 0.0034 - 0.0164 | Witteveen et al. [39] |
| Met > DBC | 0.0731 | 0.06340–0.08280 | Mo et al. [40] |
| **Year 2** |  |  |  |
| DFS > LR | 0.00217 | 0.00192–0.00242 | Gupta et al. [37] |
| DFS > Met | 0.00375 | 0.00325–0.00417 | Gupta et al. [37] |
| LR > DFS* | 0.0088 | 0.0069 – 0.0106 | Estimated based on 10% salvage rate. |
| LR > Met** | 0.01201 | 0.00940–0.01471 | Elsisi et al. [38] |
| LR > DBC | 0.01090 | 0.00830–0.02360 | Witteveen et al. [39] |
| Met > DBC | 0.0506 | 0.044–0.057 | Mo et al. [40] |
| **Year 3-5** |  |  |  |
| DFS > LR | 0.0031 | 0.00275–0.00342 | Gupta et al. [37] |
| DFS > Met | 0.0053 | 0.00475–0.00592 | Gupta et al. [37] |
| LR > DFS* | 0.00874 | 0.0069 – 0.0106 | Estimated based on 10% salvage rate. |
| LR > Met** | 0.017 | 0.014–0.02 | Elsisi et al. [38] |
| LR > DBC | 0.0097 | 0.0062–0.013 | Witteveen et al. [39] |
| Met > DBC | 0.04360 | 0.03870–0.04860 | Mo et al. [40] |
| **Year 5+** |  |  |  |
| DFS > LR | 0.0031 | 0.0028–0.0034 | Gupta et al. [37] |
| DFS > Met | 0.0053 | 0.0048–0.0059 | Gupta et al. [37] |
| LR > DFS* | 0.00874 | 0.0069 – 0.0106 | Estimated based on 10% salvage rate. |
| LR > Met** | 0.032 | 0.02598–0.03923 | Elsisi et al. [38] |
| LR > DBC | 0.0049 | 0.00001–0.0098 | Witteveen et al. [39] |
| Met > DBC | 0.02 | 0.017–0.024 | Mo et al. [40] |

For the following transition probabilities DFS > DFS, LR > LR and Met > Met, was calculated as 1 minus the sum of all outgoing transition probabilities in each row.
\*For LR > DFS, a hypothetical transition probability was assigned, as no published estimates were identified.
\*\*For LR > MET, the transition probability was calculated using a rate-based adjustment of the DFS>MET transition parameters using specified hazard ratios obtained from the stated source [36].

Transition probabilities for the 12-month regimen were used as the reference, and regimen-specific risk ratios were applied to derive the corresponding transition probabilities for the other regimens. The risk ratios for each regimen and their sources are presented in Table 3.

**Table 3:** Risk ratios of different regimens when compared to the 12-month regimen of trastuzumab.

| Name | Risk Ratio | Source |
| --- | --- | --- |
| **<u>Chemotherapy only</u>** |  |  |
| Local Recurrence | 1.667 (1.471 – 1.887) | Perez et al. [41] |
| Metastasis | 1.667 (1.471 – 1.887) |  |
| Death by Breast Cancer | 1.587 (1.370 – 1.852) |  |
| **<u>9-week regimen</u>** |  |  |
| Local Recurrence | 1.39 (1.12 – 1.72) | Joensuu et al. [10] |
| Metastasis | 1.24 (0.93 – 1.65) |  |
| Death by Breast Cancer | 1.36 (0.98 – 1.89) |  |
| **<u>6-month regimen</u>** |  |  |
| Local Recurrence | 1.07 (0.97 – 1.33) | Deng et al. [12] |
| Metastasis | 1.04 (0.82 – 1.31) |  |
| Death by Breast Cancer | 1.14 (0.99 – 1.32) |  |

| <u>9-month regimen</u> |  |  |
| --- | --- | --- |
| Local Recurrence | 1.16 (1.06 – 1.27) | Yu et al. [42] |
| Metastasis | 1.16 (1.06 – 1.27) |  |
| Death by Breast Cancer | 1.12 (0.97 – 1.30) |  |
| <u>24-month regimen</u> |  |  |
| Local Recurrence | 1.02 (0.89 – 1.17) | Cameron et al. [24] |
| Metastasis | 0.998 (0.875-1.139) |  |
| Death by Breast Cancer | 1 (0.99-1.01) |  |

Data on the health-related quality of life (HRQoL) for breast cancer patients were obtained from literature, as no published study reporting the HRQoL for breast cancer in Kenya was found. Health outcomes were measured in quality-adjusted life years (QALYs). Health utility values were assigned to each tunnel state, and these are detailed in Table 4.

**Table 4:** Health utility parameters for the tunnel states utilized in the Markov model.

| **Parameter** | **Method used** | **Base** | **Min.** | **Max.** | **Reference** |
| --- | --- | --- | --- | --- | --- |
| Diagnosis (Core Biopsy) | Testing morbidity index (TMI) | 0.84 | 0.63 | 0.93 | Swan et al. [43] |
| Surgery (Breast Conservation Surgery) | Standard Gamble (SG) | 0.76 | 0.683 | 0.827 | Songtish et al. [44] |
| Radiotherapy | EuroQol - 5 Dimension (EQ-5D) | 0.77 | 0.73 | 0.8 | Prescott et al. [45] |
| Doxorubicin + Cyclophosphamide (AC) | EQ-5D | 0.71 | 0.6 | 0.95 | Garrison et al. [46] |
| Paclitaxel + trastuzumab | EQ-5D | 0.71 | 0.6 | 0.95 | Garrison et al. [46] |
| Trastuzumab only | Time trade off (TTO) | 0.9 | 0.85 | 0.94 | Lidgren et al. [47] |
| Disease free survival (Anastrozole) in Year 1-2 | EQ-5D | 0.73 | 0.62 | 0.84 | Seferina et al. [48] |
| Disease free survival (Anastrozole) in Year 2+ | EQ-5D | 0.805 | 0.65 | 0.93 | Seferina et al. [48] |
| Diagnosis (Bone Metastasis) | TMI | 0.84 | 0.63 | 0.93 | Swan et al. [43] |
| Radiotherapy (Bone Metastasis) | TTO | 0.41 | 0 | 0.86 | Matza et al. [49] |
| Chemotherapy (AC + Zoledronic Acid) | TTO | 0.47 | 0.02 | 0.89 | Matza et al. [49] |
| Chemotherapy (Zoledronic Acid + | TTO and SG | 0.36 | 0.09 | 0.63 | Bonomi et al. [50] |
| Palliative care) |  |  |  |  |  |
| Modified Radical Mastectomy<br>(Local recurrence) | SG | 0.88 | 0.84 | 0.98 | Hayman et al. [51] |
| Chemotherapy (Capecitabine +<br>trastuzumab) | SG and TTO | 0.7 | 0.5 | 0.8 | Matter-Walstra et al.<br>[52] |
| Radiotherapy (Local recurrence) | SG | 0.61 | 0.35 | 0.87 | Kim et al. [53] |
| Disease free survival (Tamoxifen) | SG | 0.88 | 0.77 | 0.99 | Mansel et al. [54] |

#### Model outputs

Total discounted costs were calculated by multiplying the cost of each tunnel state by the number of individuals in that state for each cycle, applying the monthly discount rate for costs. The total costs for all cycles were then summed up to obtain the total discounted costs. The same approach was used to estimate the total discounted outcomes, expressed as quality adjusted life-years (QALYs). As a monthly cycle length was employed, the resulting total utilities were adjusted by a factor of 1/12.

Finally, the ICER was calculated as described in Eq. (1) by dividing the difference in total discounted costs between the intervention and reference group by the difference in their total discounted outcomes (QALYs):

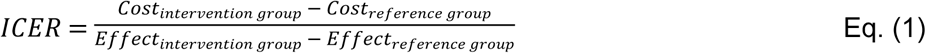

A regimen was considered to be cost-effective in the Kenyan context, if the ICER value was below the country’s willingness to pay (WTP) threshold. As no official WTP threshold existed for Kenya, we utilized a threshold of USD 1,054.80, equivalent to half of the 2022 Kenyan GDP per capita of USD 2,109.60 [55,56].

#### Model validation

Model validation followed the International Society for Pharmacoeconomics and Outcomes Research / Society for Medical Decision Making best practice guidelines on model transparency and validation, included assessment of face, internal, external and cross model validity [57]. Face validity was established through expert review of the model structure, assumptions, and clinical pathways. Internal validity was evaluated through model debugging, verification of transition matrices, and checking cost calculations. External validity was assessed by comparing model outputs with published epidemiological data, whereas cross model validity was examined through comparison with previously published models addressing similar decision problems.

#### Sensitivity analyses

Probabilistic sensitivity and deterministic (one-way) analyses were conducted using *dampack* package in R studio (build 401) [33,58]. These analyses used the minimum, maximum, base values, distribution types, means, and standard deviations of parameters, which are detailed in Tables A-F in S2_Appendix. The results of the deterministic sensitivity analysis were illustrated using tornado diagrams, while the results of the probabilistic sensitivity analysis were presented as an ICER confidence ellipse and a cost-effectiveness acceptability curve.

#### Scenario analyses

As per the recent MOU between Roche and Kenya’s Ministry of Health, implemented through the Social Health Authority (SHA), the reimbursed cost of trastuzumab was capped at KES 40,000 per cycle [20]. A scenario analysis was conducted to evaluate the impact of this pricing policy on the cost-effectiveness of the regimens. In this scenario, the trastuzumab-attributable component of all cycle costs (including local recurrence treatment) was capped at KES 40,000 in the Markov model, with other direct medical costs unchanged. The model was rerun to generate updated discounted and undiscounted costs and ICERs, which were compared to the results of the primary analysis.

A secondary scenario analysis was conducted to determine the threshold price of a vial of trastuzumab at which each trastuzumab-containing regimen would cost-effective. This analysis varied trastuzumab prices in increments of KES 500 and identified, for each regimen, the highest price at which the ICER fell below the specified WTP thresholds. These thresholds ranged from 0.5 to 3 times Kenya’s GDP per capita. The analysis aimed to inform policymakers on potential price negotiation targets for trastuzumab.

### Budget impact analysis

A budget impact analysis (BIA) estimated the financial cost of providing full treatment to all eligible patients over a five-year period for each treatment regimen. The total undiscounted cost for each regimen per year for 6100 eligible patients was obtained from the Markov model. The estimated 5-year expenditure on claims for the national insurer was calculated from data obtained from literature. NHIF’s reimbursement for patient benefits for five financial years (2020-2024) were extracted from the auditor general’s report for NHIF [59–63]. We assumed that the historic NHIF values were directly applicable to SHA, as SHA took over NHIF’s functions and maintained a similar benefits framework. The percentage share of the estimated 5-year benefit expense that each regimen would consume was calculated using the formula present in Eq (2):

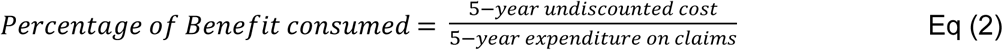

### Ethical considerations

The ethical approval for this study was obtained from the University of Nairobi/ Kenyatta National Hospital Ethics and Research Committee through the approval letter referenced P871/11/2019. Informed consent was obtained from all key informants prior to their participation. Key informants were anonymized, and all audio recordings were transcribed and subsequently deleted.

## Results

### Cost components

The cost analysis highlighted a significant difference in the cost of conventional treatment with non-biologicals, compared to targeted therapies with trastuzumab. While cost of standard chemotherapy and diagnostic procedures were relatively low, the introduction of trastuzumab drastically increased the overall cost. A single continuation cycle of paclitaxel and trastuzumab was approximately 17 times more expensive than paclitaxel alone. Furthermore, the loading dose required for trastuzumab regimens costs nearly twice as much as a subsequent continuation cycle. The cost of remission-based treatment drops by over 99% when compared to that of primary treatment. This is further detailed in Table 5.

**Table 5:** Median costs of diagnostic, treatment, and follow-up care for HER2+ breast cancer.

| Cost Description | Median cost<br>[Inter quartile range]<br>(2022, KES) | Median cost<br>[Inter quartile<br>range]<br>(2022, USD) |
| --- | --- | --- |
| Diagnosis (cost of all diagnostic tests and procedures) | 43,873.28<br>[35,821.23 - 52,723.75] | 362.59<br>[296.04 - 435.73] |
| Early-stage Radiotherapy (cost of 15 sessions) | 49,986.32<br>[32,976.26 - 74,570.73] | 413.11<br>[272.53 - 616.29] |
| Doxorubicin & cyclophosphamide (cost per cycle) | 8,120.35<br>[6,544.40 - 9,963.39] | 67.11<br>[54.09 - 82.34] |
| Paclitaxel (cost per cycle) | 9,077.96<br>[6,611.87 - 13,361.29] | 75.02<br>[54.64 - 110.42] |
| Fluorouracil, epirubicin & cyclophosphamide (cost per cycle) | 15,448.57<br>[12,973.18 - 18,298.03] | 127.67<br>[107.22 - 151.22] |
| Loading dose for Paclitaxel & trastuzumab (cost per cycle) | 152,324.76<br>[90,170.96 - 244,272.48] | 1,258.88<br>[745.21 - 2,018.78] |
| Continuation dose for Paclitaxel & trastuzumab (cost per cycle) | 81,661.06<br>[50,752.41 - 128,194.53] | 674.88<br>[419.44 - 1,059.46] |
| Trastuzumab (cost per cycle) | 76,580.09<br>[45,355.08 - 121,967.12] | 632.89<br>[374.84 - 1,007.99] |
| Disease free survival with anastrozole for the first 2 years (cost per cycle) | 1,181.96<br>[650.55 - 2,107.01] | 9.77<br>[5.38 - 17.41] |
| Disease free survival with anastrozole for year 2-5 (cost per cycle) | 1,106.96<br>[576.16 - 1,931.25] | 9.15<br>[4.76 - 15.96] |
| Radiotherapy for Local recurrence/ bone metastasis (5 sessions) | 28,085.03<br>[20,581.68 - 38,538.25] | 232.11<br>[170.10 - 318.50] |
| Local recurrence treatment with capecitabine and loading dose of trastuzumab (cost per cycle) | 164,882.47<br>[105,849.19 - 257,804.24] | 1,362.67<br>[874.79 - 2,130.61] |
| Local recurrence treatment with capecitabine and continuation dose of trastuzumab (cost per cycle) | 95,735.67<br>[65,455.69 - 144,151.25] | 791.20<br>[540.96 - 1,191.33] |
| Metastasis treatment with chemotherapy and zoledronic acid (cost per cycle) | 10,901.86<br>[8,770.77 - 13,788.21] | 90.10<br>[72.49 - 113.95] |
| Metastasis treatment with palliative care and zoledronic acid (cost per cycle) | 29,428.29<br>[20,774.53 - 40,836.48] | 243.21<br>[171.69 - 337.49] |

### Cost trajectory of treatment over 5 years

The annual costs associated with different treatment regimens over the 5-year period are shown in Fig2. Costs for most treatment groups (Chemotherapy only, 9-week, 6- and 9-month) were highest in Year 1, reflecting on the primary treatment phase. The costs of the longer duration trastuzumab regimens peaked in the second year (12- and 24-month), where majority of the trastuzumab related costs occurred. Costs declined progressively from Year 2 onward as primary treatment phases ended for most regimens and patients received treatment for remission. By the end of the third year, the costs for all strategies except the 24-month regimen were minimal.

**Fig2:**
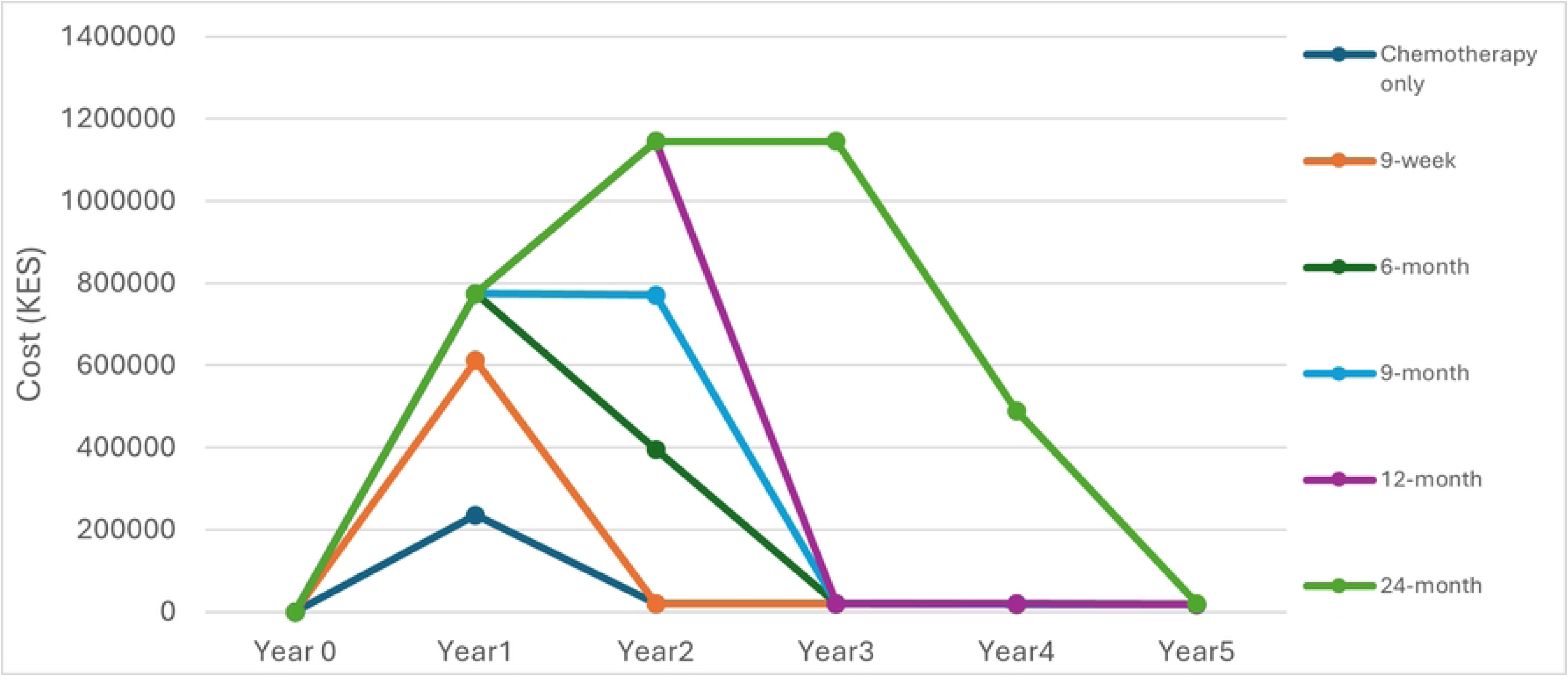
Annual costs per treatment regimen over a five-year costing time horizon

### Cost-utility analysis

#### Base-case analysis

The regimen with the lowest ICER was the 9-week regimen at 3,230 USD/QALY. The 6-, 9-, 12-, and 24-month regimens had ICER values ranging from 4,046 to 9,846 USD/QALY. Notably, the 9-month regimen was dominated (as shown in Fig3), as the incremental costs exceeded the incremental clinical benefits. The discounted and incremental costs and QALYs, and the ICERs for each regimen are detailed in Table 8.

**Fig3:**
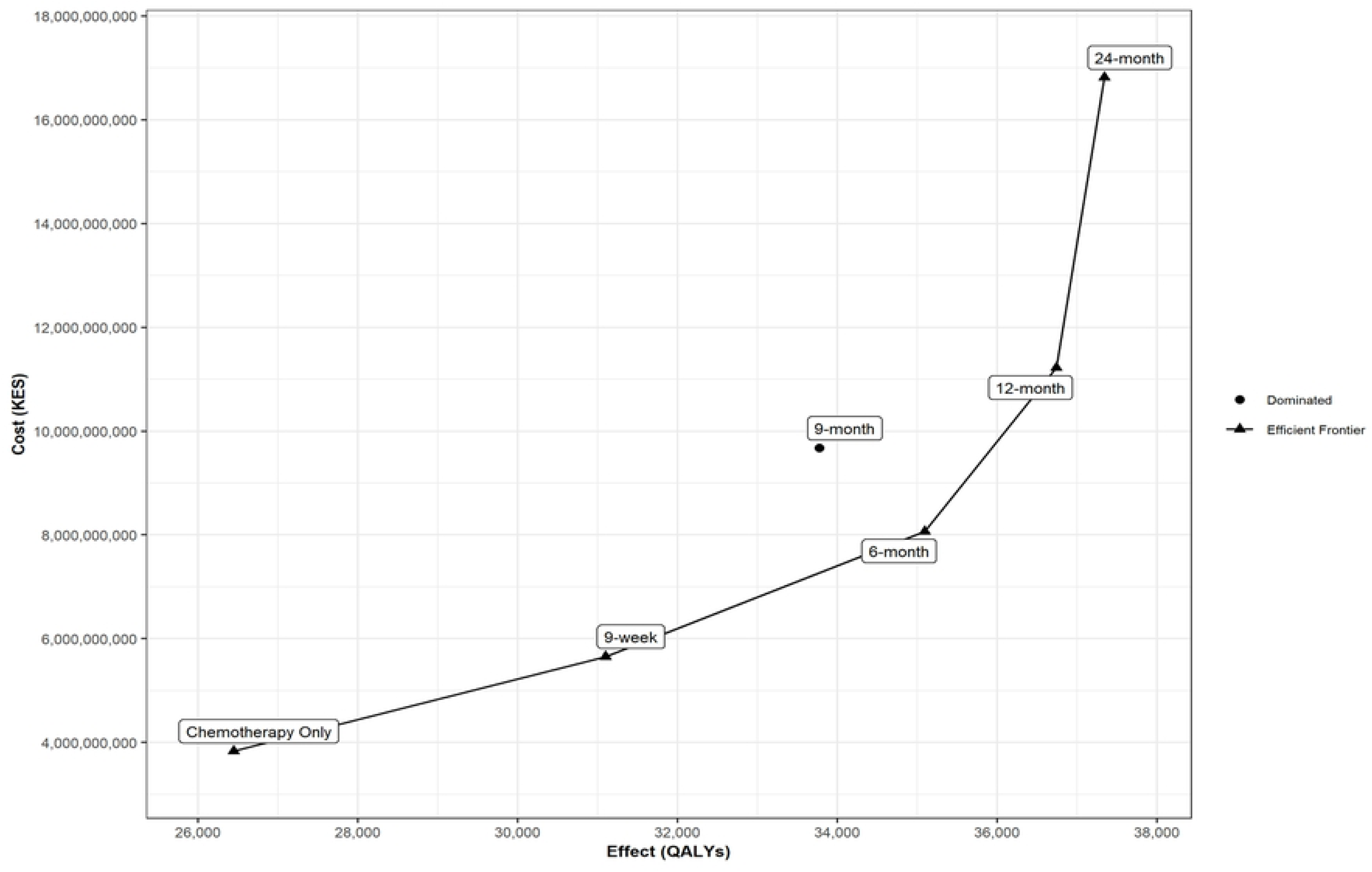
Cost-effectiveness plane depicting cost and effectiveness of the different regimens

**Table 8:** Discounted total and incremental costs and QALYs, and ICERs for each regimen.

| Parameter | Total Discounted Cost<br>(2022, KES) | Total Discounted<br>Cost<br>(2022, USD) | Total<br>Discounted<br>QALYs |
| --- | --- | --- | --- |
| Chemotherapy only | 3,836,756,760 | 31,708,734 | 26,444 |
| 9-week regimen | 5,655,676,462 | 46,741,128 | 31,098 |
| 6-month regimen | 8,068,623,202 | 66,682,836 | 35,089 |
| 9-month regimen | 9,675,912,420 | 79,966,218 | 33,775 |
| 12-month regimen | 11,220,962,574 | 92,735,228 | 36,737 |
| 24-month regimen | 16,817,230,476 | 138,985,376 | 37,339 |
| Differences | Incremental Cost<br>(2022, KES) | Incremental Cost<br>(2022, USD) | Incremental<br>QALYs |
| Chemotherapy only | Reference | Reference | Reference |
| 9-week regimen | 1,818,919,702 | 15,032,394 | 4,654 |
| 6-month regimen | 4,231,866,441 | 34,974,103 | 8,645 |
| 9-month regimen | 5,839,155,660 | 48,257,485 | 7,331 |
| 12-month regimen | 7,384,205,813 | 61,026,494 | 10,293 |

| 24-month regimen | 12,980,473,716 | 107,276,642 | 10,895 |
| --- | --- | --- | --- |
| ICER | ICER (KES/QALY) | ICER (USD/QALY) |  |
| Chemotherapy only | Reference | Reference |  |
| 9-week regimen | 390,868 | 3,230 |  |
| 6-month regimen | 489,524 | 4,046 |  |
| 9-month regimen | 796,546 | 6,583 |  |
| 12-month regimen | 717,418 | 5,929 |  |
| 24-month regimen | 1,191,415 | 9,846 |  |

#### Sensitivity analysis

The cost-effectiveness acceptability curve in Fig4 illustrates how the probability that each regimen is cost-effective changes depending on the WTP threshold. Chemotherapy only starts with the highest probability of being cost-effective but declines sharply. At 1 times GDP per capita (KES 255,260; USD 2,110), the 9-week and 6-month regimens emerge as the most competitive alternatives. At 2 times GDP per capita (KES 510,520; USD 4,219), no strategy clearly dominates, with chemotherapy only, 9-week, and 6-month each holding approximately 30% probability. Beyond 3 times GDP per capita (KES 765,780; USD 6,329), the 6-month regimen becomes the most acceptable cost-effective strategy, plateauing at approximately 40%. The 9-month regimen remained consistently near zero across all thresholds.

**Fig4:**
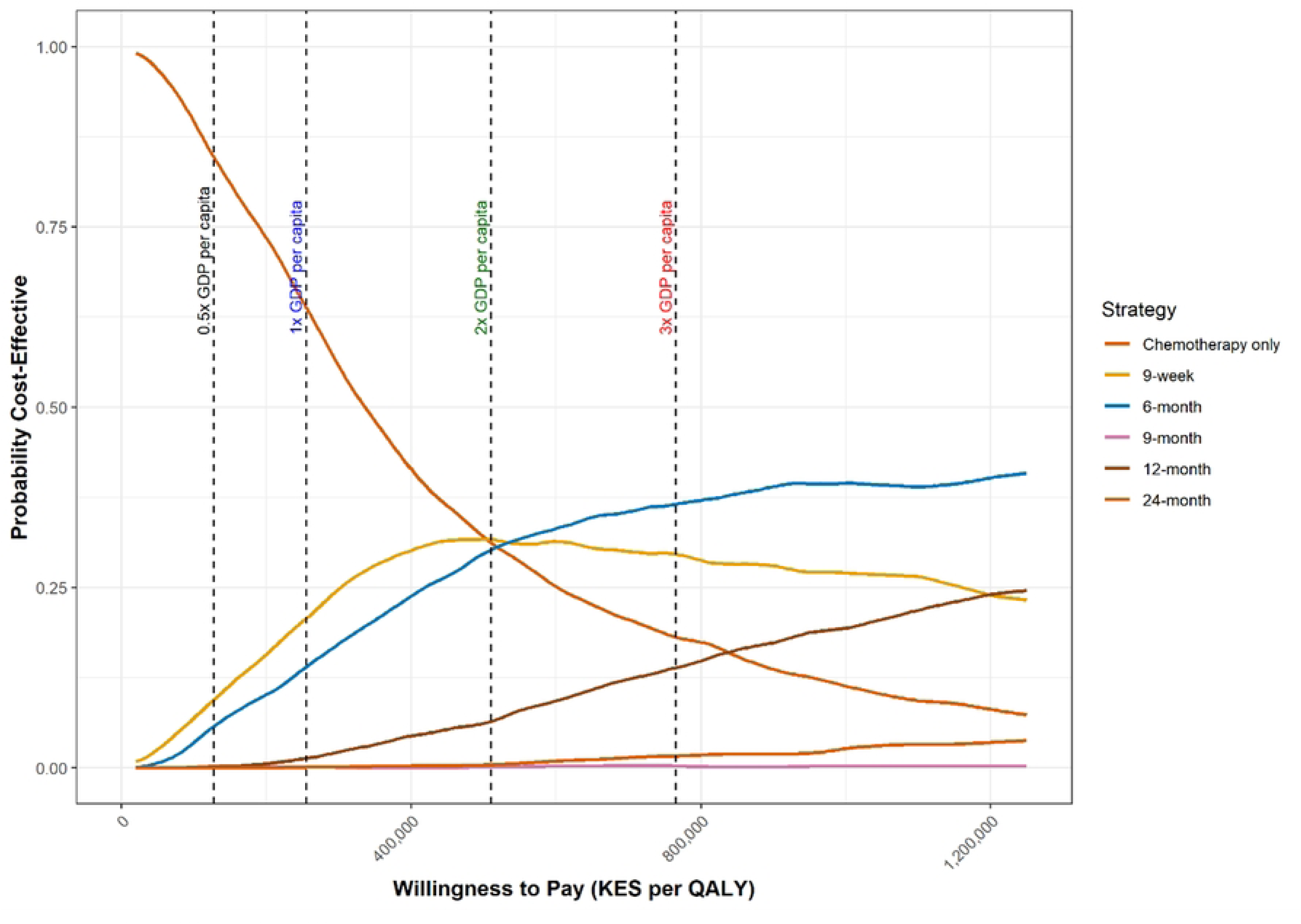
Cost-effectiveness acceptability curve

The cost-effectiveness scatter plot shown in Fig5 highlights substantial overlap in the probabilistic simulations across the different trastuzumab regimens. The longer duration regimens (12- and 24-month) produce PSA iterations that encompass the point estimates of the shorter duration regimen (9-week). This suggests that under certain conditions for cost and effectiveness, the longer duration regimens could have the same cost effectiveness as the 9-week regimen.

**Fig5:**
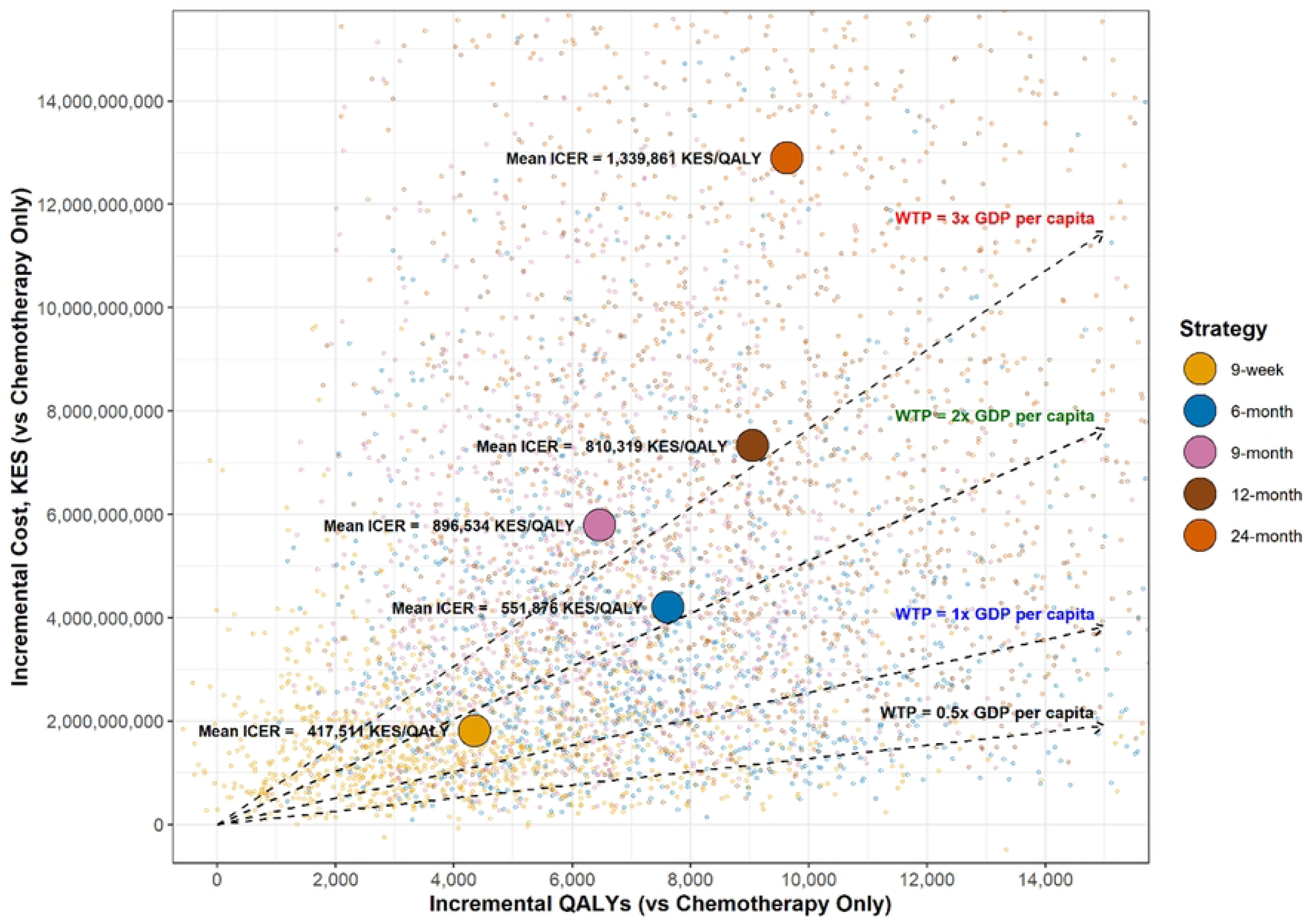
Cost-effectiveness plane.

The tornado diagrams in Fig6 show which model parameters the ICERs for the 9-week and 12-month group were most sensitive to, with S1_Fig illustrating the tornado diagrams for the remaining regimens. Across five of the six regimens, the price of trastuzumab was the parameter to which the ICER was most sensitive. The exception was the 9-week regimen, where the risk ratio for metastasis was the most influential parameter. Additional parameters that notably influenced the ICER across regimens included the discount rate, the ratio of the discount rate applied to costs versus utilities, and the quantity of trastuzumab consumed.

**Fig6:**
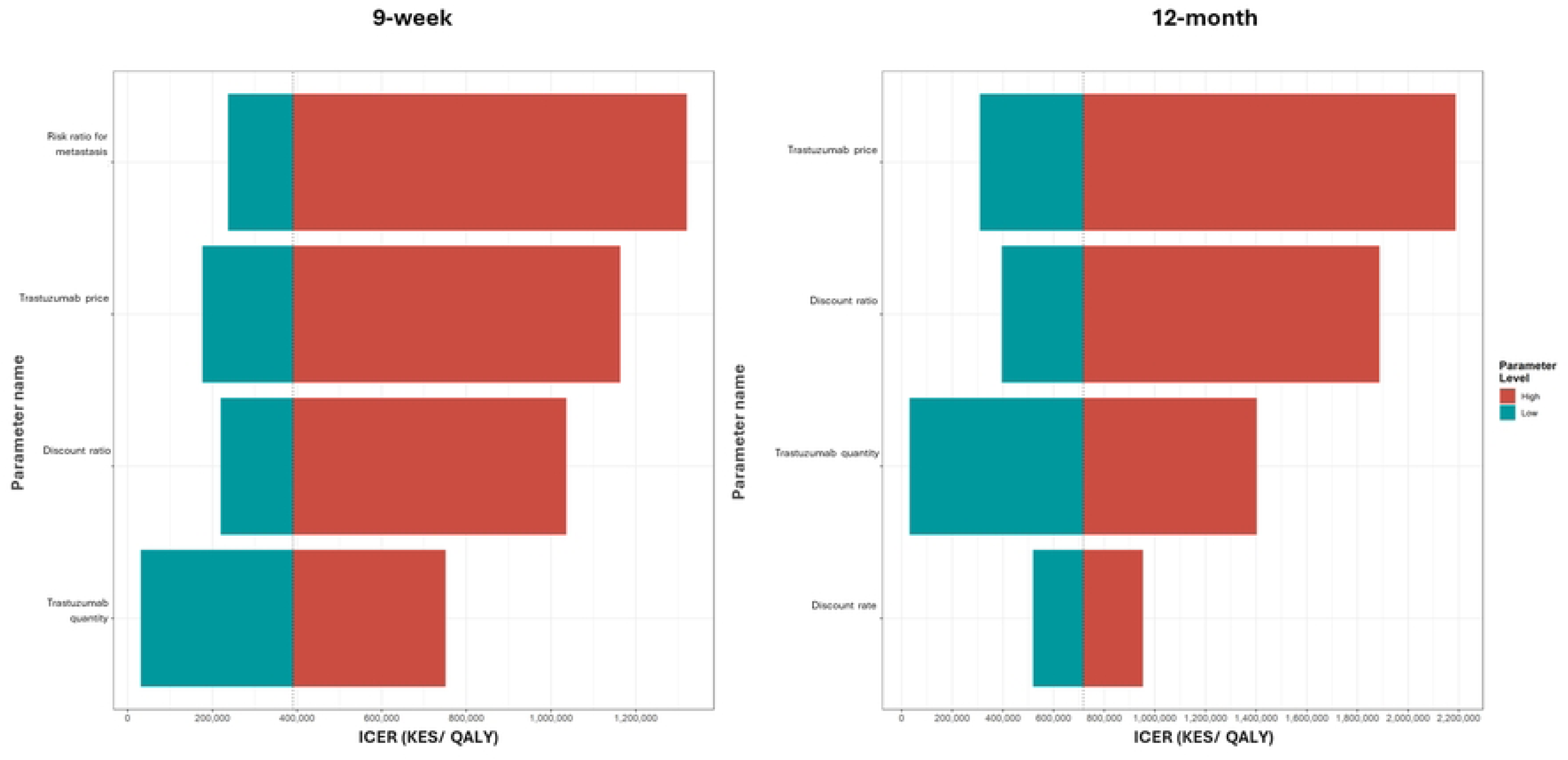
Tornado diagrams for incremental cost-effectiveness ratio of the 9-week and 12-month regimen

### Scenario analysis

#### Trastuzumab reimbursement cap

In the scenario analysis incorporating the KES 40,000 per-cycle cap on trastuzumab reimbursement costs, lead to substantial reductions in total costs and ICERs across all trastuzumab-containing regimens when compared to the results of the primary analysis. The reduction in total cost was most significant in the 24-month group, given the higher number of trastuzumab cycles. In contrast, the 9-week group had the greatest reduction in ICER (61% reduction), as it had the fewest treatment cycles with trastuzumab (n=3), resulting in trastuzumab-related costs making up a large proportion of the total costs and increased its sensitivity to the fixed per-cycle reimbursement cap. Notably, even the chemotherapy only reference regimen experienced a modest cost reduction (11%), as patients transitioning to local recurrence received capped trastuzumab-based salvage therapy. The results are detailed in Table 9.

**Table 9:** Scenario analysis: Impact of per cycle reimbursement cap on trastuzumab.

| Strategy | Scenario |  |  | Percentage reduction |  |  |
| --- | --- | --- | --- | --- | --- | --- |
|  | Undiscounted costs (USD) | Discounted costs (USD) | ICER (USD/QALY) | Undiscounted costs (%) | Discounted costs (%) | Incremental costs and ICER (%) |
| Chemotherapy only | 43,129,977 | 28,385,371 | Reference | 10.95% | 10.48% | Reference |
| 9-week | 52,279,329 | 34,243,625 | 1,259 | 24.35% | 26.74% | 61.03% |
| 6-month | 64,531,979 | 43,195,493 | 1,713 | 31.96% | 35.22% | 57.65% |
| 9-month | 72,262,507 | 49,721,598 | 2,911 | 34.91% | 37.82% | 55.79% |
| 12-month | 80,988,983 | 55,766,836 | 2,660 | 36.98% | 39.86% | 55.13% |
| 24-month | 111,007,349 | 77,888,604 | 4,544 | 41.96% | 43.96% | 53.85% |

#### Threshold price of trastuzumab

The second scenario analysis examined the threshold price of trastuzumab per vial at which each regimen remains cost-effective across four WTP thresholds. At a WTP of 0.5 times GDP per capita, the 9-week regimen had the highest threshold price per vial of KES 23,500 (USD 194), while the 24-month regimen had the lowest of KES 5,500 (USD 45). As the WTP threshold increases, threshold prices rose across all regimens, with the 9-week, 6-month, and 12-month regimens reaching the current market price per vial at 2 times, 2 times, and 3 times GDP per capita WTP thresholds respectively. The 9- and 24-month regimen did not reach the current market price per vial at any threshold.

### Budget-impact analysis

The estimated 5-year expenditure on claims from 2020 to 2024 was just over USD 2.5 billion. The projected cost for the estimated 6100 patients over the 5 years in the nine-week group was just under USD 30.8 million (lowest of all the trastuzumab-containing regimens). It would account for 1.2% of the forecasted SHA expenditure. For the remaining regimens, total costs increased in line with the duration of trastuzumab treatment, with their projected share of expenditure ranging from 2.0% to 4.0%. The 12-month group, which is the current recommended regimen, would account for 2.8% of the expenditure. Based on the annual budget impact threshold’s proposed by Pichon-Riviera et al. [64] for Kenya, all treatment regimens exceeded the very-high budget impact threshold of USD 0.98 million. This is summarized in Table 10.

**Table 10:** Threshold analysis of trastuzumab vial prices.

| Strategy | Willingness to pay threshold | Threshold trastuzumab price per vial, KES (USD) |
| --- | --- | --- |
| 9-week | 0.5x GDP | 23500 (194) |
|  | 1x GDP | 55000 (455) |
|  | 2x GDP | Current price* |
|  | 3x GDP | Current price* |
| 6-month | 0.5x GDP | 20500 (169) |
|  | 1x GDP | 44500 (368) |
|  | 2x GDP | Current price* |
|  | 3x GDP | Current price* |
| 9-month | 0.5x GDP | 10500 (87) |
|  | 1x GDP | 25500 (211) |
|  | 2x GDP | 55500 (459) |
|  | 3x GDP | 85000 (702) |
| 12-month | 0.5x GDP | 12000 (99) |
|  | 1x GDP | 28500 (236) |
|  | 2x GDP | 62000 (512) |
|  | 3x GDP | Current price* |
| 24-month | 0.5x GDP | 5500 (45) |
|  | 1x GDP | 15500 (128) |
|  | 2x GDP | 35500 (293) |
|  | 3x GDP | 55500 (459) |

\*Current price: KES 88,995 (USD 735)

**Table 10:** Impact of the different regimens on the social health authority budget.

|  | **Chemotherapy**<br>**Only**<br>**KES, (USD)**<br>**millions** | **9-week**<br>**KES,**<br>**(USD)**<br>**millions** | **6-month**<br>**KES,**<br>**(USD)**<br>**millions** | **9-month**<br>**KES,**<br>**(USD)**<br>**millions** | **12-month**<br>**KES,**<br>**(USD)**<br>**millions** | **24-month**<br>**KES,**<br>**(USD)**<br>**millions** |
| --- | --- | --- | --- | --- | --- | --- |
| **Year 1** | 206.99<br>(1.71) | 276.23<br>(2.28) | 349.29<br>(2.89) | 349.10<br>(2.89) | 349.34<br>(2.89) | 349.37<br>(2.89) |
| **Year 2** | 338.65<br>(2.80) | 753.58<br>(6.23) | 1,284.16<br>(10.61) | 1,481.27<br>(12.24) | 1,553.58<br>(12.84) | 1,553.55<br>(12.84) |
| **Year 3** | 416.15<br>(3.44) | 824.04<br>(6.81) | 1,415.74<br>(11.70) | 1,814.48<br>(15.00) | 2,213.09<br>(18.29) | 2,772.23<br>(22.91) |
| **Year 4** | 499.08<br>(4.12) | 901.12<br>(7.45) | 1,485.40<br>(12.28) | 1,887.74<br>(15.60) | 2,281.49<br>(18.86) | 3,710.73<br>(30.67) |
| **Year 5** | 573.32<br>(4.74) | 972.44<br>(8.04) | 1,551.16<br>(12.82) | 1,956.53<br>(16.17) | 2,346.78<br>(19.39) | 3,860.88<br>(31.91) |
| **5-year total cost** | 2,034.19 (16.81) | 3,727.42<br>(30.81) | 6,085.75<br>(50.30) | 7,489.12<br>(61.89) | 8,744.26<br>(72.27) | 12,246.75<br>(101.21) |
| **Percentage of**<br>**forecasted 5-**<br>**year SHA**<br>**expenditure**<br>**utilized** | 0.65% | 1.20% | 1.96% | 2.41% | 2.82% | 3.94% |
| **Budget impact**<br>**classification\*** | Very high | Very<br>high | Very high | Very high | Very high | Very high |

Values presented in millions of KES with USD equivalents in parentheses.
\*Budget impact categories based on Pichon-Riviera et al. et al. for Kenya

## Discussion

The question this study aimed to answer was whether trastuzumab was cost-effective and affordable in Kenya and, if so, what would be the ideal duration for its use. We compared the cost and effectiveness of various trastuzumab-based regimens against a reference group that did not contain trastuzumab. With Kenya not having a specific WTP threshold, we adopted a value of USD 1054.80 [55,56]. At the stated threshold, none of the trastuzumab-based regimens were cost-effective in Kenya. However, at higher thresholds of 2 times the GDP per capita (USD 4,219), the 9-week and 6-month regimens are cost-effective. Given the severity of breast cancer and its impact on quality of life, higher WTP thresholds may be justifiable, as noted by Schurer et al. [65], which would support the adoption of shorter trastuzumab regimens in the Kenyan context. Studies done in the Philippines, Iran, and across multiple countries in sub-Saharan Africa also showed trastuzumab not to be cost-effective [22,66,67], whereas studies from Thailand and Vietnam that used thresholds of 3 times and 1 times their national GDP per capita respectively, reported the contrary [68,69].

The total cost of treatment varied substantially across the different regimens. The 5-year treatment cost with the chemotherapy only regimen was USD2,584, whereas the cost rose significantly once trastuzumab was introduced, ranging from USD5,694 (9-week) up to USD29,535 (24-month). When comparing our results with the literature, the total cost for the chemotherapy only regimen (AC-T) in our study was higher than that reported by Gitonga et al. [29]. The lower costs in their study could be explained by the exclusion of the cost of radiotherapy planning fees and cost of subsequent hormonal treatment during the remission phase, and use of fewer radiotherapy sessions (4 v/s 15) during treatment, when compared to our analysis. For the trastuzumab regimens, the study by Gershon et al. [22] utilized lower treatment cost for the 12-month regimen in Kenya; however, it is important to note that the study utilized the same cost across all countries, which likely explains the difference from our Kenya-specific estimates.

The study showed that the 9-week regimen had the lowest ICER and was the most affordable regimen for Kenya. Similar results were seen in India with the 9-week regimen having the lowest ICER [37]. Although the chemotherapeutic agents in the 9-week regimen differ from those recommended in Kenyan guidelines, policymakers could still consider it a viable option due to its affordability. Conversely, the 9-month regimen emerged as a dominated strategy; this was largely due to use of efficacy estimates derived from a systematic review that modeled its comparative performance against the 12-month regimen [42], due to the lack of long-term clinical data. The currently recommended 12-month regimen had a significantly higher ICER (84%) and would consume more than double the projected expenditure of the national insurer, when compared with the 9-week regimen. The shorter duration of the 9-week schedule could also enhance patient adherence and lessen the strain on the healthcare system, further reinforcing its value in resource limited settings.

When placed in a global context, the ICERs reported in our study were lower than those reported from most middle-income countries [22,66,70,71], though higher than those reported in Thailand and India [37,69], and comparable to recent findings from Indonesia [68]. These differences are likely attributable to several methodological factors. First, this study adopted a provider perspective, whereas some comparable studies employed a societal perspective [36,70], which typically captures a broader range of costs and may therefore yield higher ICERs. Second, variations in the specific cost items included and their unit prices across settings reflect differences in local healthcare costs and pricing structures. Third, differences in the structure and underlying assumptions of the Markov models used across studies, including transition probabilities, time horizons, discount rates, and the definition of health states, further contributed to the observed variation in ICERs across settings.

Sensitivity analysis demonstrated that the quantity and price of trastuzumab were the parameters to which the ICERs were most sensitive, a finding consistent with similar studies in the literature [22,72,73]. On the quantity side, interventions such as vial sharing and good dispensing practices could help reduce trastuzumab consumption per patient, while for the drug price, cost-reduction strategies including price negotiations with suppliers and the introduction of biosimilars could meaningfully lower drug acquisition costs. Notable, the Kenyan government has already made strides in this front with an MOU signed with the manufacturer of the drug (Roche) to provide the drug at subsidized prices [20]. For the 9-week regimen specifically, the risk ratio for metastasis was also identified as a highly sensitive parameter, highlighting the need for locally generated evidence on the effectiveness of this regimen in the Kenyan context.

Our scenario analysis clearly highlights that the MOU has significantly reduced overall treatment costs as well as the ICERs across all regimens. However, it should be noted that the subsidized price established through the MOU may not reflect the true economic cost of trastuzumab, and therefore the ICERs reported under this scenario should be interpreted with caution rather than taken as a direct reflection of the drug’s cost-effectiveness at its market value. Interventions that aid in early diagnosis and treatment initiation, such as breast cancer screening, patient navigation programs and training of healthcare workers, can further help improve health outcomes and reduce the ICER. Furthermore, our threshold analysis revealed that at a WTP threshold of 0.5 times the GDP per capita, the 9-week regimen supported the highest vial price (USD 194), suggesting it is the most economically resilient strategy for the Kenyan context. These estimates can serve as a practical reference for policymakers and procurers when negotiating target trastuzumab prices with manufacturers, helping to anchor discussions around what constitutes an affordable and cost-effective price point for the Kenyan setting.

The BIA projected that the 9-week regimen would consume 1.2% of the forecasted SHA budget if all diagnosed patients were enrolled, representing a modest increase over chemotherapy alone. Although the BIA showed that even the 9-week regimen exceeds the “very-high” budget impact threshold of USD 0.98 million for Kenya [64], a 1-2% increase in the budget could potentially be obtained through strategic resource mobilization. Such an approach aligns with SHA’s policy direction concerning social insurance, were higher earners pay higher premiums to help cushion the poor [74]. Another implementation option would be to use a phase-wise matrix, which would award full coverage to vulnerable groups at the start, followed by an annual increase in the proportion of patients covered for this treatment. This strategy would effectively mitigate the initial fiscal shock and ensure more sustainable integration into the national insurance framework.

Some of the strengths of this study include being the first economic evaluation of trastuzumab for HER2-positive breast cancer in Kenya that incorporates local cost data, providing context-specific evidence to guide policy. The study used tunnel states in the Markov model to give a more accurate reflection of a patient’s journey. The study also analysed multiple trastuzumab-based regimens to provide a broad policy perspective. Where possible, real-world data were used, offering a more realistic representation of clinical practice and treatment outcomes. The study also included a policy-relevant budget impact and scenario analyses to help highlight the benefits of price negotiations and affordability. Furthermore, this is the first economic evaluation to analyze five different trastuzumab regimens simultaneously.

The study also had some limitations. Health utility values were drawn from systematic reviews due to the absence of Kenyan-specific data, and although sensitivity analyses were undertaken to adjust for these assumptions, there is still a clear need for local quality-of-life surveys. We utilized data from literature to estimate transition probabilities, as Kenyan studies reported unrealistically low mortality rates, largely due to loss to follow-up, poor adherence, and treatment abandonment. In addition, we assumed 100% coverage for interventions in the BIA, whereas in practice, coverage increases gradually over time, meaning that the budget impact over the next five years may be overestimated compared to what would be realized in real-world implementation. Finally, some costing inputs used in the sensitivity analysis were based on estimates obtained from KIIs, which may have been subject to response bias.

It is unlikely that in a resource-limited country like Kenya, any trastuzumab-containing regimen would be cost-effective. Most African countries do not readily use trastuzumab for treatment due to the high cost of treatment, which was highlighted by Vanderpuye et al. (10), who stated that out-of-pocket payment for trastuzumab is affordable for less than 20% of patients in sub-Saharan Africa. The cost of treatment with the currently recommended 12-month regimen, estimated at approximately USD 16,400, is unaffordable for a majority of Kenyans. Given that the average annual income, represented by the gross national income per capita, is only USD 1,864 [75], the cost of a full course of treatment equates to more than eight years of income for an average citizen. Policymakers should consider prioritizing the 9-week regimen, which requires less than half the budget of the 12-month regimen, and represents that most cost-effective trastuzumab-based option.

## Conclusion

This study demonstrates that while trastuzumab-based regimens are not cost-effective at Kenya’s adopted willingness-to-pay threshold, the 9-week regimen represents the most economically viable option, with the lowest ICER and a budget impact of 1.2% of forecasted SHA expenditure. The currently recommended 12-month regimen is unaffordable for the majority of Kenyans, costing the equivalent of more than eight years of average income. Price negotiations, as evidenced by the MOU with Roche, biosimilar uptake, and improved dispensing practices are critical to reducing costs and improving the value proposition of trastuzumab in Kenya. A phased incorporation of the 9-week regimen into the national benefits package, prioritizing vulnerable groups, represents the most fiscally sustainable pathway to expanding access. This study provides the first Kenya-specific economic evidence on multiple trastuzumab regimens simultaneously, offering a robust foundation for policy decisions on the adoption and financing of trastuzumab within the national health insurance framework.

## Data Availability

All data underlying the findings of this study are available within the manuscript and its supporting information files.

## Supporting Information Captions

### S1_Appendix. Tunnel States by Health State and Treatment Regimen

The tunnel state structure for each health state, outlining the order of treatment and care costs applied by cycle number

### S2_ Appendix. Input parameters for model and sensitivity analysis

All input parameters used in the Markov model, including costs, transition probabilities, utility values, and risk ratios.

**S1_Fig. Tornado diagrams for incremental cost-effectiveness ratio of the 6-, 9-, and 24-month trastuzumab regimens.**

Tornado diagrams illustrating the parameters to which the ICER was most sensitive for the 6-, 9-, and 24-month regimens

